# Self-versus caregiver-reported apathy across neurological disorders

**DOI:** 10.1101/2025.02.14.25322300

**Authors:** Sijia Zhao, Anna Scholcz, Matthew A. Rouse, Verena S. Klar, Akke Ganse-Dumrath, Sofia Toniolo, M. John Broulidakis, Matthew A. Lambon Ralph, James B. Rowe, Peter Garrard, Sian Thompson, Sarosh R Irani, Sanjay Manohar, Masud Husain

## Abstract

Apathy is a prevalent and persistent neuropsychiatric syndrome across many neurological disorders, significantly impacting on both patients and caregivers. We systematically quantified discrepancies between self- and caregiver-reported apathy in 335 patients with a variety of diagnoses, frontotemporal dementia (behavioural variant and semantic dementia subtypes), Parkinson’s disease, Parkinson’s disease dementia, dementia with Lewy bodies, Alzheimer’s disease dementia, mild cognitive impairment, small vessel cerebrovascular disease, subjective cognitive decline and autoimmune encephalitis.

Using the Apathy-Motivation Index (AMI) and its analogous caregiver version (AMI-CG), we found that caregiver-reported apathy consistently exceeded self-reported levels across all conditions. Moreover, self-reported apathy accounted for only 14.1% of the variance in caregiver ratings. This apathy reporting discrepancy was most pronounced in conditions associated with impaired insight, such as behavioural variant frontotemporal dementia, and was significantly correlated with cognitive impairment. Deficits in memory and fluency explained an additional 11.2% of the variance in caregiver-reported apathy, highlighting their crucial role in goal-directed behaviour. Specifically, executive function deficits (e.g., indexed by fluency) and memory impairments may contribute to behavioural inertia or recall of it.

These findings highlight the need to integrate patient and caregiver perspectives in apathy assessments, especially for conditions with prominent cognitive impairment. To improve diagnostic accuracy and deepen our understanding of apathy across neurological disorders, we emphasise the need of standardised apathy assessment tools tailored to individuals with cognitive deficits. Understanding the cognitive mechanisms underpinning discordant apathy reporting in dementia might help to inform targeted clinical interventions and reduce caregiver burden.

## Introduction

Apathy is defined as a syndrome of reduced goal-directed behaviours, including social activity or emotional responsiveness, associated with reduced motivation affecting and significantly impacting on everyday life.^1^ It is increasingly recognised as a prevalent and persistent problem across neurodegenerative disorders,^2–7^ and associated with functional impairment, independent of disease severity and other covariates (e.g. age, sex, and depression).^8–15^ The syndrome presents significant challenges for both patients and caregivers. Apathetic individuals often rely on caregivers to initiate activities they previously used to perform independently.^16^ Apathy may be associated with lower quality of life, although it may also protect against distress in the presence of impairments.^14,17–21^ At the same time, caregivers often misinterpret apathy as a voluntary choice by the patient not to engage, which makes it one of the most distressing neuropsychiatric symptoms they report.^14,17–21^ Accurate assessment of apathy is therefore crucial for effective management, but this can be challenging due to potential discrepancies between patient self-report and caregiver perspectives.^22,23^ However, while apathy is prevalent across all dementias, the extent to which self- or caregiver reports captures it differs between disorders, highlighting the need for a systematic evaluation of this discrepancy.

Previous research, using qualitative interviews^24–26^ or established questionnaires such as the Lille Apathy Rating Scale (LARS)^27^ and Apathy Evaluation Scale (AES)^28^, has examined the discrepancy in apathy between patients and caregivers in Alzheimer’s disease dementia (AD), frontotemporal dementia (FTD) and Parkinson’s disease (PD).^27,29–31^ These studies revealed that caregivers generally perceived patients as more apathetic than the patients self-reported, and that the correlation between caregiver and patient assessments is low.^24–27,29–32^ This may reflect caregiver over-estimation, potentially due to increased burden,^27,30^ patient underestimation,^24–26^ or patient violations of the assumptions underlying the use of rating scales.^33^ Underestimates from self-reporting has been linked with cognitive impairment in dementia, which may represent anosognosia.^34–38^ Impaired awareness of one’s own deficits has been associated with greater apathy severity in AD, without corresponding increases in anxiety or depression.^36,37,39,40^ A recent report showed that this phenomenon was particularly prominent in frontotemporal dementia (FTD).^40^

Here we used the Apathy Motivation Index (AMI), a validated self-report measure of apathy,^41^ and its caregiver-report counterpart (AMI-CG),^42^ to investigate the discrepancy between self- and caregiver-reported apathy across ten neurological conditions: behavioural variant FTD (bvFTD), semantic dementia (SD), PD, PD dementia (PDD), dementia with Lewy bodies (DLB), AD, Mild Cognitive Impairement (MCI), small vessel cerebrovascular disease (SVD), autoimmune encephalitis (AIE) and Subjective Cognitive Decline (SCD). Apathy is extremely common in bvFTD,^15,32,43–45^ with most studies reporting greater than 70% patients affected at some point in their illness.^46^ In PDD and DLB, apathy also affects over 70% of individuals and is associated with increased caregiver burden and functional impairment.^47,48^ Here we systematically compare self- and care-giver-reported apathy across this range of conditions using analogous scales.

We hypothesise that cognitive impairment may contribute to self and caregiver-reported apathy discrepancy across these neurological conditions. Specifically, we predict a strong relationship between cognitive performance and the degree of discrepancy in apathy ratings. Furthermore, we anticipate greater discrepancies in conditions associated with prominent insight deficits, such as bvFTD. Confirmation of these hypotheses would underscore the importance of educating caregivers about the influence of cognitive impairment and specific diagnoses (e.g. bvFTD) on apathy awareness. This knowledge could help alleviate caregiver burden and facilitate the development of targeted support strategies.

## Materials and methods

### Participants

This study included 335 patients with various neurological conditions and their respective care-givers. The patient cohort comprised the following ten neurological conditions: bvFTD (N = 44), SD (N = 15), SVD (N = 24), PD (N = 58), PDD (N = 19), DLB (N = 18), AD (N = 54), MCI (N = 7), and SCD (N = 43) (**Table 1**).

**Table 1.** Demographics and clinical characteristics of participants. For continuous variables (age, length of patient-caregiver relationship, etc.), the mean and standard deviation (SD) are reported, along with the number of participants for whom this information was available (indicated in parentheses). Abbreviations: bvFTD = behavioural variant frontotemporal dementia; SD = semantic dementia; SVD = small vessel disease; AIE = autoimmune encephalitis; PD = Parkinson’s disease; PDD = Parkinson’s disease dementia; DLB = dementia with Lewy bodies; AD = Alzheimer’s disease; MCI = mild cognitive impairment; SCD = subjective cognitive decline; HC = healthy controls. Note: “All patients” refers to all diagnostic groups combined, excluding healthy controls. * = data missing. ACE-III = Addenbrooke’s Cognitive Examination-III. **AMI-SR = Apathy-Motivation Index Self-reported version. AMI-CG = Apathy-Motivation Index caregiver version**

| Diagnosis | N | Gender | Caregiver Gender | Age | Caregiver Age | Length of Relationship | Education | ACE Total | AMI-SR Total | AMI-CG Total |
| --- | --- | --- | --- | --- | --- | --- | --- | --- | --- | --- |
| bvFTD | 43 | F13 M30 | F15 M7 (N=22) | 62.95 (9.55, N=40) | 62.37 (10.93, N=19) | 44.55 (13.38, N=22) | 12.50 (2.41, N=14) | 66.11 (15.82, N=38) | 1.42 (0.79, N=43) | 2.63 (0.74, N=43) |
| SD | 16 | F11 M5 | * | 64.69 (7.43, N=16) | * | * | * | 59.81 (14.46, N=16) | 1.22 (0.35, N=16) | 1.69 (0.76, N=16) |
| SVD | 24 | F4 M20 | F17 M3 (N=20) | 70.08 (7.56, N=24) | 66.11 (7.29, N=18) | 42.91 (14.55, N=22) | 12.29 (2.81, N=21) | 80.50 (18.43, N=22) | 1.38 (0.50, N=24) | 1.77 (0.66, N=24) |
| AIE | 53 | F18 M35 | F20 M4 (N=24) | 63.38 (13.45, N=53) | 54.29 (16.40, N=14) | 36.10 (16.19, N=51) | 12.80 (3.34, N=50) | 90.85 (6.45, N=47) | 1.39 (0.57, N=53) | 1.56 (0.68, N=53) |
| PD | 58 | F23 M35 | F6 M1 (N=7) | 68.47 (7.63, N=58) | 59.43 (17.00, N=7) | 42.04 (15.20, N=57) | 15.75 (4.10, N=53) | 94.40 (3.53, N=53) | 1.41 (0.44, N=58) | 1.58 (0.60, N=58) |
| PDD | 19 | F4 M15 | F10 M1 (N=11) | 71.05 (7.55, N=19) | 66.67 (12.85, N=9) | 38.89 (14.52, N=18) | 12.50 (3.06, N=12) | 73.21 (11.97, N=19) | 1.60 (0.55, N=19) | 2.11 (0.66, N=19) |
| DLB | 18 | F4 M13 | F15 M3 (N=18) | 69.24 (7.23, N=17) | 61.41 (12.57, N=17) | 38.83 (17.54, N=18) | 14.40 (4.75, N=15) | 75.61 (15.81, N=18) | 1.48 (0.62, N=18) | 2.03 (0.65, N=18) |
| AD | 54 | F24 M30 | F16 M5 (N=21) | 69.33 (9.39, N=54) | 60.33 (14.87, N=12) | 42.49 (14.29, N=53) | 14.21 (3.64, N=47) | 67.82 (16.61, N=51) | 1.58 (0.56, N=54) | 1.86 (0.68, N=54) |
| MCI | 7 | F6 M1 | F2 M1 (N=3) | 64.57 (6.90, N=7) | 63.00 (0.00, N=1) | 42.14 (10.33, N=7) | 12.57 (2.88, N=7) | 85.29 (5.59, N=7) | 1.26 (0.22, N=7) | 1.72 (0.59, N=7) |
| SCD | 43 | F20 M23 | F8 M3 (N=11) | 57.02 (9.98, N=43) | 61.40 (8.11, N=5) | 30.74 (14.67, N=43) | 14.95 (3.66, N=38) | 91.88 (7.89, N=43) | 1.66 (0.48, N=43) | 1.63 (0.68, N=43) |
| HC | 19 | F11 M8 | * | 64.26 (6.71, N=19) | * | * | * | 97.16 (1.57, N=19) | 1.12 (0.31, N=19) | 0.92 (0.46, N=19) |
| All patients | 335 | F127 M207 | F109 M28 (N=137) | 65.68 (10.39, N=331) | 61.66 (12.68, N=102) | 39.24 (15.41, N=292) | 13.99 (3.77, N=258) | 80.48 (17.04, N=314) | 1.47 (0.56, N=335) | 1.84 (0.75, N=335) |

The majority of patients (N = 247) were recruited from the Cognitive Disorders Clinic at John Radcliffe Hospital, Oxford, UK. All AIE patients (N = 53) were recruited from Autoimmune Neurology Clinic. 35 patients (all with FTD, either bvFTD or SD) were recruited from specialist dementia clinics in Addenbrooke’s Hospital, Cambridge, UK (N = 31) and St George’s Hospital, London, UK (N = 4). Nineteen older healthy controls and their partners were also included, with most of these dyads (18/19) recruited in Cambridge. The questionnaire data were collected between 12 July 2016 and 6 May 2024. All diagnoses were made by experienced neurologists according to consensus clinical diagnostic critieria.

Demographic information, including patient and caregiver age, gender, relationship to the patient, and length of the caregiver-patient relationship, was collected for most participants. A summary of demographic characteristics for each cohort is presented in **Table 1**.

Caregiver relationship to the patient was recorded for 298 of the Oxford patients. Of these, 237 were spouses or partners, 29 were children, 21 were siblings or other family members, and 11 were friends. No professional caregivers were included in this sample. The majority of caregivers were female (79.6% of 137 caregivers with gender recorded).

To be included in the study, caregivers were required to know the patient well, defined as either being a spouse/partner (79.5% of the sample) or having known the patient for at least 3 years. On average, caregivers had known the patients for 39.2 years (SD = 15.4 years; see **Table 1** for cohort-wise statistics).

### Measures

**Apathy:** All patients completed the Apathy-Motivation Index Self-reported version (here referred as AMI-SR)^41^, and their caregivers completed the caregiver version (AMI-CG)^42^.

- AMI-SR: This 18-item self-report questionnaire assesses apathy across three domains: Behavioural Activation, Social Motivation, and Emotional Sensitivity. Participants respond using a 5-point Likert scale. Item scores are averaged to yield subscale and total scores, with higher scores indicating greater apathy (range 0–4). A cut-off score of ≥ 1.91 (1 SD above the mean of a healthy population) was used to identify apathy.^41^
- AMI-CG: This measure uses the same items and response options as the AMI, with wording adapted to reflect a third-person perspective. ^42^ Developed from the original AMI, this measure covers the same three domains of apathy: Behavioural, Social and Emotional Apathy. It is scored in the same way, with higher scores indicating greater apathy (range 0–4). Since the AMI-CG is adapted from the self-reported AMI questionnaire and contains identical item content, but no caregiver-specific cutoff score has been established, we applied the same cutoff (≥ 1.91) as used for AMI-SR.

**Depression and Anhedonia:** Patients recruited in Oxford (N=300) also completed the following:

- **Snaith-Hamilton Pleasure Scale (SHAPS)**^49^: This 14-item self-report questionnaire aims to assess hedonic tone across four domains: interest/pastimes, social interaction, sensory experience, and food/drink. A four-point response scale was used, with higher scores indicating greater anhedonia. Following Franken, Rassin, & Muris (2007), a four-point scoring system was used to increase data dispersion for correlation analyses. A cut-off score of ≥ 22.3, derived from a recent meta-analysis, was used to identify clinically significant anhedonia.^50^ Data were available for 298 patients.
- **Geriatric Depression Scale Short Form (GDS)**^51^: This 15-item self-report screening tool aims to assess depressive symptoms in older adults excluding somatic symptoms, providing a more robust measure in individuals with neurodegenerative illnesses. A two-point response scale (yes/no) was used, with scores ranging from 0–15; higher scores indicate more severe depression. An established cut-off score of ≥ 5 was used to identify possible depression.^52,53^ To minimise overlap with apathy, a GDS Depression subscale score was calculated, excluding the three *prima facie* apathy-related items: “Have you dropped many of your activities and interests?”, “Do you prefer to stay at home, rather than going out and doing new things?”, and “Do you feel full of energy?”. This adjustment did not affect the overall conclusions of the study. Data were available for 294 patients.

**Cognitive Function:** Patients recruited in Oxford also completed the Addenbrooke’s Cognitive Examination-III (ACE-III)^54^ in person (N=279), and 35 patients recruited in Cambridge completed the Addenbrooke’s Cognitive Examination-Revised (ACE-R).^55^ They are very similar and both assessed cognitive function across five domains: attention, memory, verbal fluency, language, and visuospatial skills. Scores range from 0–100, with lower scores indicating greater cognitive impairment. Both ACE-III and ACE-R scores > 88/100 are generally considered normal.^56^ The total score (ACE Total) and individual subscores for each domain (ACE attention, ACE memory, ACE fluency, ACE language and ACE visuospatial) were reported here.

### Statistical analysis

All analyses were performed using MATLAB (version R2024b), R statistical software (version 4.3.3)^57^ and R-based statistical software JASP^58^.

Exploratory factor analysis (EFA) was conducted using R package psych.^59^ The Kaiser–Meyer–Olkin (KMO) measure of sampling adequacy was first computed to assess whether the sample was appropriate for factor analysis. To determine the optimal number of factors, Horn’s Parallel Analysis was conducted with 2,000 iterations. EFA was then conducted using Promax rotation, which allows for inter-factor correlations. This choice was based on prior knowledge that the different subdomains of apathy measured by the AMI are interrelated. While this approach reduces factor orthogonality, we found no difference in factor structure when using Varimax rotation. Therefore, we chose to report the EFA results using Promax rotation. Reliability of the AMI-CG was assessed using Cronbach’s alpha, computed in JASP.

All bivariate and partial correlations were performed using Spearman’s rank correlation. Following Cohen (1988), correlation coefficients ≥ 0.1, ≥ 0.3, and ≥ 0.5 were interpreted as representing weak, moderate, and strong relationships, respectively. Differences in Spearman correlation coefficients were tested using the procedures for testing statistical differences between correlations using the implementation in the R package cocor.^60^

All p values reported are two-tailed. Group comparisons were performed using paired t-tests for paired data. For between-group comparisons with unequal sample sizes, continuous variables were compared using the Mann–Whitney U test, and categorical variables were compared using the χ^2^ test. Effect sizes for between-group comparisons were estimated using rank-biserial correlation. All p-values reported in correlation matrix figures are adjusted for multiple comparisons using Bonferroni correction.

### Ethics statement

The study was performed in accordance with the Declaration of Helsinki. All participants, including caregivers, gave written informed consent. Ethical approval was granted by the University of Oxford ethics committee (IRAS ID: 248379, Ethics Approval Reference: 18/SC/0448). For the Cambridge and London participants, ethical approval was granted by the National Re-search Ethics Service’s East of England Cambridge Central Committee (IRAS ID: 252986).

### Data availability

The data supporting the findings of this study are available from the corresponding author, upon reasonable request.

## Results

### Prevalence of apathy, depression and anhedonia in neurological disorders

We began by examining the prevalence of self-reported apathy and related neuropsychiatric syndromes—depression and anhedonia—in our patient cohorts. Patients were classified as apathetic based on their AMI-SR total score, depressed based on their GDS total score, and anhedonic based on their SHAPS total score (see Methods for cutoffs). The prevalence of each syndrome and their overlaps are illustrated in **Figure 1**. Overall, across the 292 patients who completed all three questionnaires, 21.6% self-reported clinically significant apathy, 45.5% met criteria for depression self-reported significant depression symptoms and 47.6% exceeded the threshold for anhedonia. Considerable overlap was observed between these syndromes, with 11.3% of the 293 patients exceeding the threshold for all three. The prevalence of each syndrome within each diagnostic cohort is presented in **Table 2**.

**Figure 1:**
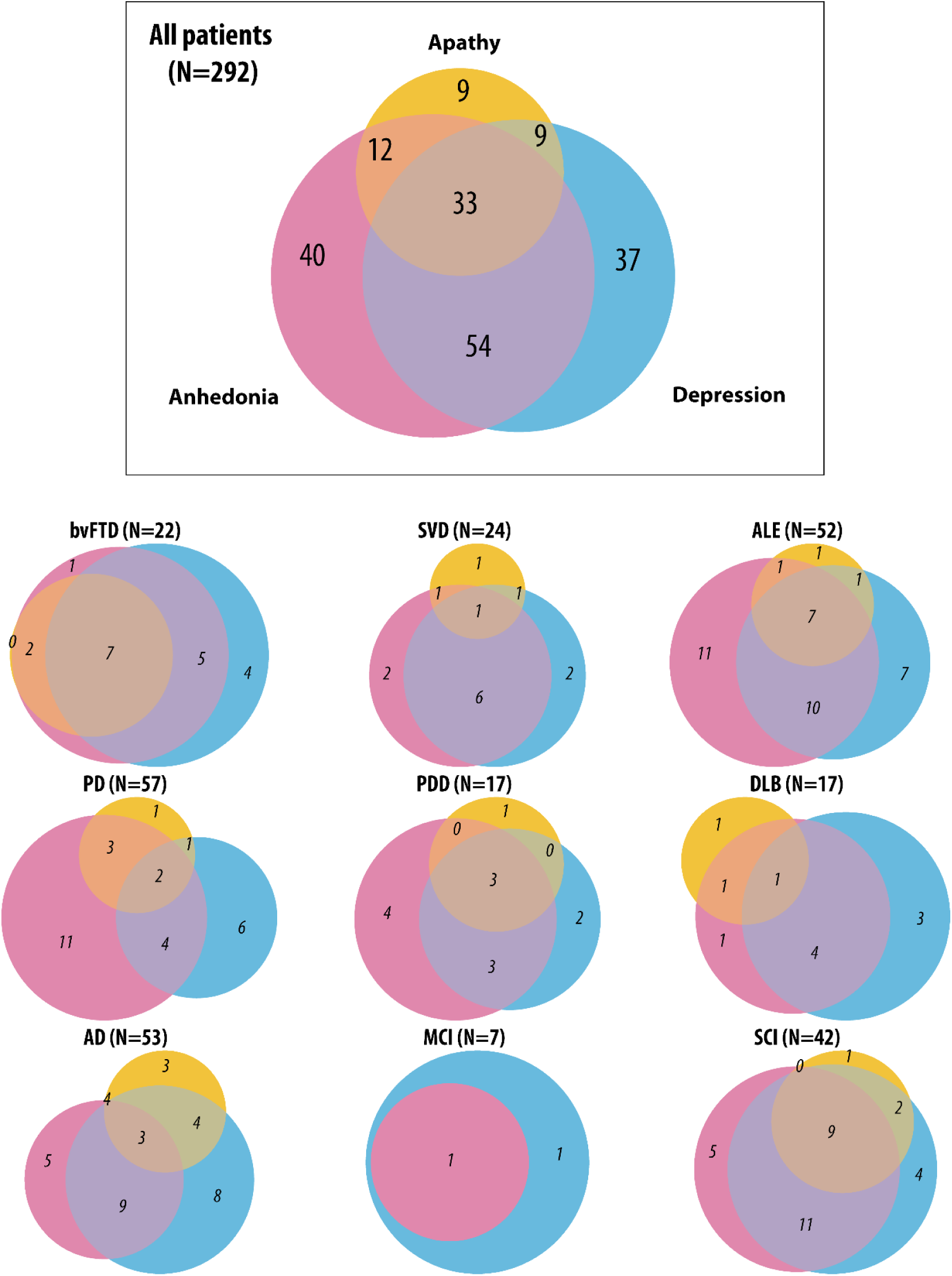
Prevalence of Self-Reported Apathy, Depression, and Anhedonia. Venn diagram illustrating the overlap between self-reported apathy, depression, and anhedonia Yellow, pink, and blue circles show apathy, anhedonia, and depression, respectively. See Methods for the cut-offs. Note that the SD group was not included here aswe only had one patient who completed all AMI, SHAPS, and GDS questionnaires from this group. The details of the prevalence (including who had none of the syndromes) can be found in Table 2.

**Table 2:** Prevalence of self-reported apathy, depression and anhedonia. The percentage was computed based on all patients (N=292) in that diagnostic group who completed all three questionnaires: AMI for apathy, GDS for depression, and SHAPS for anhedonia. ’No ADA’ refers to the proportion of individuals who did not experience apathy, depression, or anhedonia. ’ADA’ indicates those who experienced all three conditions—apathy, depression, and anhedonia.Only one patient in the Semantic Dementia (SD) group completed all three questionnaires (AMI, GDS, and SHAPS), so the prevalence for this group is not reported here.

| Group | N | SR Apathy | Depression | Anhedonia | No ADA | ADA | Apathy & Depression | Apathy & Anhedonia | Depression & Anhedonia | Pure Apathy | Pure Depression | Pure Anhedonia |
| --- | --- | --- | --- | --- | --- | --- | --- | --- | --- | --- | --- | --- |
| bvFTD | 22 | 40.9% | 72.7% | 68.2% | 13.6% | 31.8% | 0.0% | 9.1% | 22.7% | 0.0% | 18.2% | 4.5% |
| SVD | 24 | 16.7% | 41.7% | 41.7% | 41.7% | 4.2% | 4.2% | 4.2% | 25.0% | 4.2% | 8.3% | 8.3% |
| AIE | 52 | 19.2% | 48.1% | 55.8% | 26.9% | 13.5% | 1.9% | 1.9% | 19.2% | 1.9% | 13.5% | 21.2% |
| PD | 57 | 12.3% | 22.8% | 35.1% | 50.9% | 3.5% | 1.8% | 5.3% | 7.0% | 1.8% | 10.5% | 19.3% |
| PDD | 17 | 23.5% | 47.1% | 58.8% | 23.5% | 17.6% | 0.0% | 0.0% | 17.6% | 5.9% | 11.8% | 23.5% |
| DLB | 17 | 17.6% | 47.1% | 41.2% | 35.3% | 5.9% | 0.0% | 5.9% | 23.5% | 5.9% | 17.6% | 5.9% |
| AD | 53 | 26.4% | 45.3% | 39.6% | 32.1% | 5.7% | 7.5% | 7.5% | 17.0% | 5.7% | 15.1% | 9.4% |
| MCI | 7 | 0.0% | 28.6% | 14.3% | 71.4% | 0.0% | 0.0% | 0.0% | 14.3% | 0.0% | 14.3% | 0.0% |
| SCD | 42 | 28.6% | 61.9% | 59.5% | 23.8% | 21.4% | 4.8% | 0.0% | 26.2% | 2.4% | 9.5% | 11.9% |
| All patients | 292 | 21.6% | 45.5% | 47.6% | 33.6% | 11.3% | 3.1% | 4.1% | 18.5% | 3.1% | 12.7% | 13.7% |

Focusing specifically on apathy, we compared the prevalence based on self-report and caregiver report in the 335 patients with both AMI-SR and AMI-CG data (**Table 3**). While 20.9% of patients self-reported apathy, the prevalence based on caregiver report was substantially higher, at 43.9% (**Table 3** and **Figure 2**). Although some agreement was observed between self- and caregiver report, discrepancies were evident in all neurological conditions except for SVD, where all patients identified as apathetic by self-report were also considered apathetic by their caregivers (**Figure 2**). Across all conditions, the prevalence of apathy was consistently higher based on caregiver report than on self-report.

**Figure 2:**
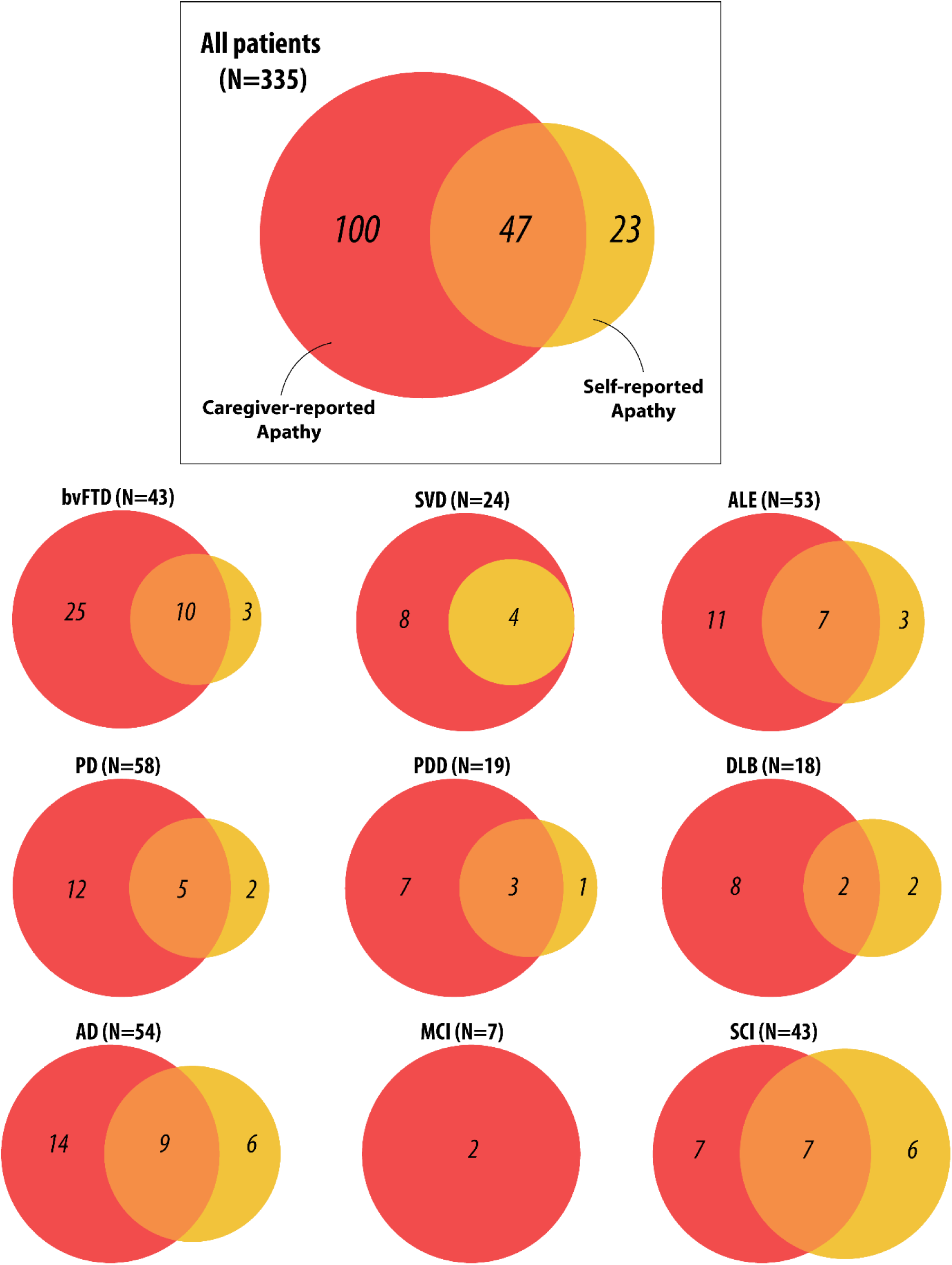
Discrepancy in apathy determined by caregiver report and self-report. Orange circle shows apathy determined by caregiver-report AMI and yellow circle shows apathy determined by self-reported AMI, using the established cutoff of 1.91 for both AMI and AMI-CG Total scores. In the SD group, no individuals were classified as apathetic based on self-reported AMI, whereas 6 out of 16 individuals were classified as apathetic based on caregiver-reported AMI.

**Table 3:** Prevalence of self-reported versus caregiver-reported supra-threshold apathy across all diagnostic groups. This table includes all individuals who completed both the AMI-SR and AMI-CG, resulting in a larger sample size compared to Table 2, which only includes patients who also completed the GDS and SHAPS.

| Group | N | Self-reported Apathy | Caregiver-reported Apathy |
| --- | --- | --- | --- |
| bvFTD | 43 | 30.2% | 81.4% |
| SD | 16 | 0.0% | 37.5% |
| SVD | 24 | 16.7% | 50.0% |
| AIE | 53 | 18.9% | 34.0% |
| PD | 58 | 12.1% | 29.3% |
| PDD | 19 | 21.1% | 52.6% |
| DLB | 18 | 22.2% | 55.6% |
| AD | 54 | 27.8% | 42.6% |
| MCI | 7 | 0.0% | 28.6% |
| SCI | 43 | 30.2% | 32.6% |
| **All patients** | **335** | **20.9%** | **43.9%** |
| HC | 19 | 0.0% | 5.3% |

When investigating apathy, depression, and anhedonia prevlance using caregiver-reported AMI, we found a big increase in apathy prevalence in all groups (**Table 4**). In summary, 292 patients across ten neurological conditions had only 27.1% free from all three syndromes; in bvFTD, only 9.1% were free from these three syndromes. Apathy was present in 41.4% of all patients, depression in 45.5%, and anhedonia in 47.6%, and 17.8% exhibited all three syndromes concurrently. Only 9.6% of patients presented with pure apathy, 10.6% with pure depression, and 8.9% with pure anhedonia, highlighting that nearly one in five patients experience these conditions in isolation.

**Table 4:** Prevalence of caregiver-reported apathy, self-reported depression and self-reported anhedonia. The percentage was computed based on all patients (N=292) in that diagnostic group who completed all three questionnaires: AMI for apathy, GDS for depression, and SHAPS for anhedonia. ’No ADA’ refers to the proportion of individuals who did not experience apathy, depression, or anhedonia. ’ADA’ indicates those who experienced all three conditions—apathy, depression, and anhedonia.Only one patient in the Semantic Dementia (SD) group completed all three questionnaires (AMI, GDS, and SHAPS), so the prevalence for this group is not reported here.

| Group | N | CG Apathy | Depression | Anhedonia | No ADA | ADA | Apathy & Depression | Apathy & Anhedonia | Depression & Anhedonia | Pure Apathy | Pure Depression | Pure Anhedonia |
| --- | --- | --- | --- | --- | --- | --- | --- | --- | --- | --- | --- | --- |
| bvFTD | 22 | 72.7% | 72.7% | 68.2% | 9.1% | 45.5% | 13.6% | 9.1% | 9.1% | 4.5% | 4.5% | 4.5% |
| SVD | 24 | 50.0% | 41.7% | 41.7% | 29.2% | 20.8% | 4.2% | 8.3% | 8.3% | 16.7% | 8.3% | 4.2% |
| AIE | 52 | 34.6% | 48.1% | 55.8% | 23.1% | 15.4% | 3.8% | 9.6% | 17.3% | 5.8% | 11.5% | 13.5% |
| PD | 57 | 29.8% | 22.8% | 35.1% | 47.4% | 10.5% | 1.8% | 12.3% | 0.0% | 5.3% | 10.5% | 12.3% |
| PDD | 17 | 52.9% | 47.1% | 58.8% | 17.6% | 29.4% | 11.8% | 0.0% | 5.9% | 11.8% | 0.0% | 23.5% |
| DLB | 17 | 52.9% | 47.1% | 41.2% | 23.5% | 23.5% | 5.9% | 5.9% | 5.9% | 17.6% | 11.8% | 5.9% |
| AD | 53 | 43.4% | 45.3% | 39.6% | 20.8% | 9.4% | 5.7% | 11.3% | 13.2% | 17.0% | 17.0% | 5.7% |
| MCI | 7 | 28.6% | 28.6% | 14.3% | 42.9% | 0.0% | 0.0% | 0.0% | 14.3% | 28.6% | 14.3% | 0.0% |
| SCD | 42 | 33.3% | 61.9% | 59.5% | 23.8% | 19.0% | 4.8% | 7.1% | 28.6% | 2.4% | 9.5% | 4.8% |
| All patients | 292 | 41.4% | 45.5% | 47.6% | 27.1% | 17.8% | 5.1% | 8.9% | 12.0% | 9.6% | 10.6% | 8.9% |

### Factor structure and reliability of the AMI-CG

The three-factor structure of the AMI, previously identified in self-reported measures in healthy individuals^41^ and caregivers of AD and PD patients^42^, was replicated in in our diverse clinical sample. Exploratory factor analysis (EFA) of AMI-SR (**Figure 3**, left) and AMI-CG (**Figure 3**, right) confirmed the presence of three distinct factors of apathy: Behavioural Inactivation, Social Disengagment, and Emotional Insensitivity.

**Figure 3:**
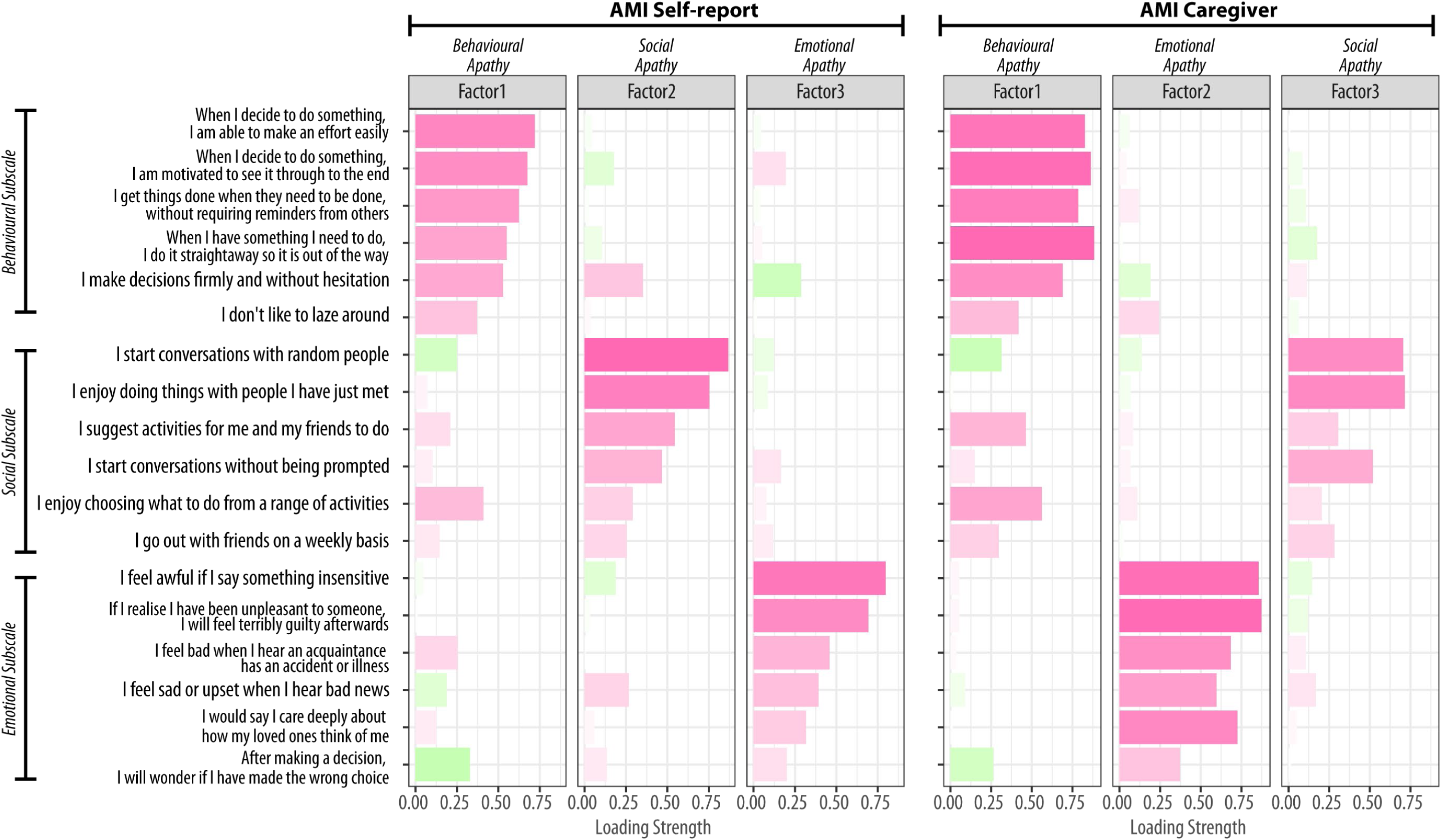
Factor structures for AMI Caregiver and AMI Self-report (N=335). (Left) Results of the exploratory factor analysis for the 18-item AMI reported by patients themselves are shown for the three factors. The item label shows the original question used in the AMI-SR. The order of items is arbitrary. Pink bars indicate positive loadings, and green bars indicate negative loadings. Factors 1, 2, and 3 primarily load on questions from the three apathy subdomains of the AMI: Behavioural Apathy, Social Apathy, and Emotional Apathy. This is consistent with the AMI-SR data from the prior studies in healthy population.^41^ (Right) The caregiver’s reports also assessed the same three apathy subdomains, but with a different factor order: Behavioural Apathy, Emotional Apathy, and Social Apathy. This structure, including the factor order, is consistent with the AMI-CG data from the prior study.^42^ Both exploratory factor analyses used parallel analysis to decide the number of factors. The factors appear non-orthogonal because Promax rotation was applied, allowing for inter-factor correlations. This choice was based on prior evidence that the subdomains of apathy are interrelated.

The KMO measure was 0.91, confirming the sampling adequacy for factor analysis. Horn’s Parallel Analysis further supported the retention of three factors. EFA yielded a three-factor model that adequately fit the data (χ2(102) = 229.4, *p* < 0.001), explaining 51.0% of the variance. The model demonstrated good fit indices (RMSEA = 0.063, 90% CI 0.053–0.074; SRMR = 0.032; TLI = 0.92; CFI = 0.95; BIC = -354.9). Note that here EFA was conducted using Promax rotation, which allows for inter-factor correlations, based on prior evidence that the subdomains of apathy measure by the AMI are interrelated.^41,42^ While this approach *reduces* factor orthogonality, the factor structure remained consistent when using Varimax rotation, confirming that the identified factors reflect meaningful constructs rather than an artifact of rotation choice. Therefore, we report the EFA results using Promax rotation.

The factor structure of the original AMI-SR was also confirmed in the AMI-CG, with Factor 1 (Behavioural Apathy), Factor 2 (Emotional Apathy), and Factor 3 (Social Apathy) loading onto corresponding items of the Behavioural Activation, Emotional Sensitivity, and Social Motivation subscales, respectively (Figure 1B). The factors were intercorrelated (r_Behaviour-Emotional_= 0.50, r_Behaviour-Social_ = 0.51, r_Emotional-Social_ = 0.46).

The reliability and construct validity of the AMI-CG were also examined. Cronbach’s alpha for the AMI-CG total score indicated good internal reliability (α_overall_ = 0.89, 95% CI 0.87–0.91). Internal consistency was good for the Behavioural Apathy (α = 0.87) and Emotional Apathy (α = 0.83) subscales, and acceptable for the Social Apathy subscale (α = 0.74).

### Discrepancy between self and caregiver reports on apathy

Despite both patients and caregivers reporting on the same apathy items, only a moderate correlation was observed between the total AMI-SR and AMI-CG scores (ρ = 0.37, *p* < 0.001, N = 335). Correlations within each subdomain were also significant but moderate, with the correlation found for Social Apathy (ρ = 0.40), Behavioural Apathy (ρ = 0.34) and Emotional Apathy (ρ = 0.30), all p < 0.001 (see **Figure 4**). There were no significant differences in the strength of these correlations.

**Figure 4:**
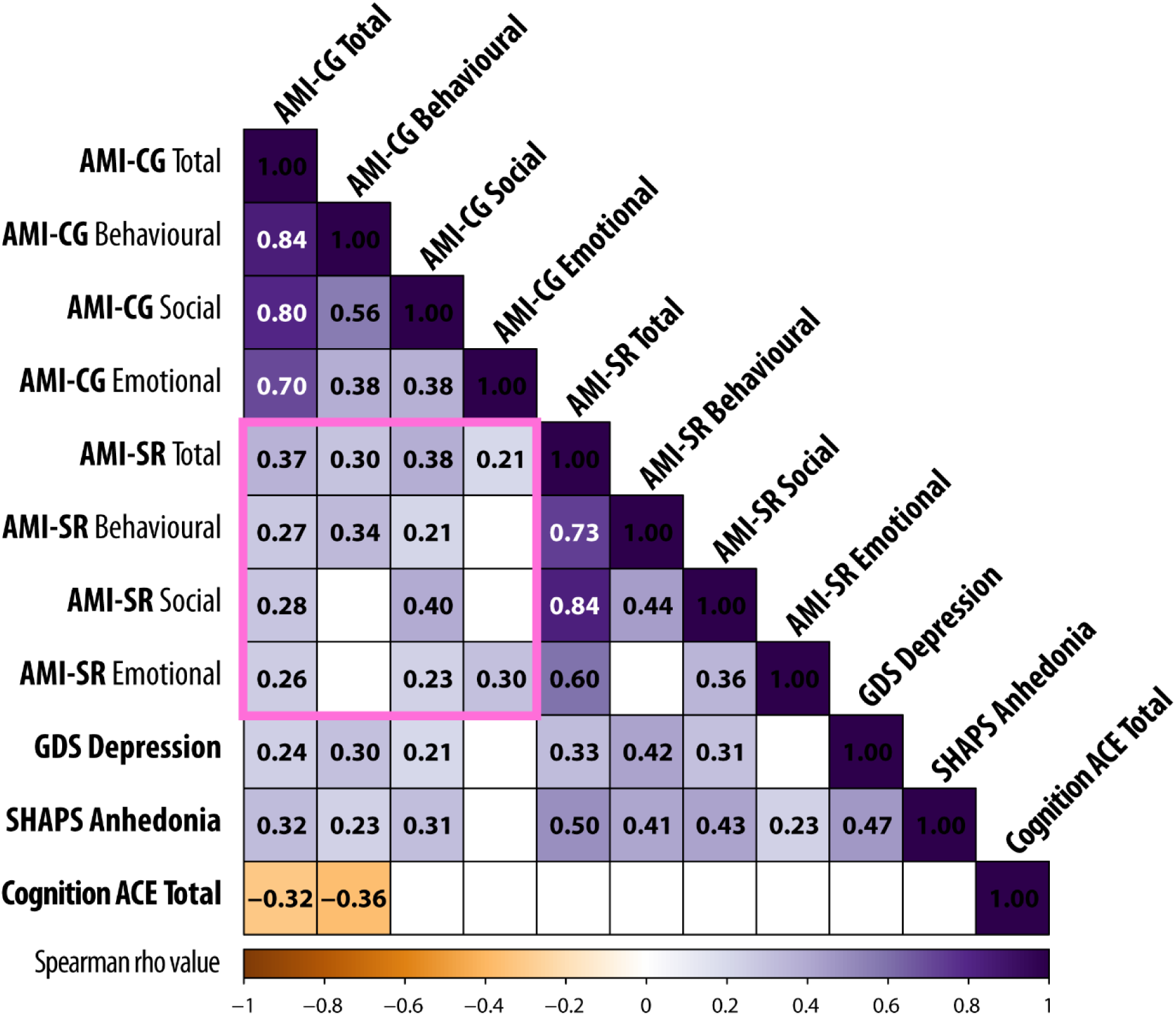
Correlation matrix for AMI-CG and self-report measures. Correlation matrix for AMI-CG total and subscale scores with self-reported AMI, GDS, and SHAPS scores, along with ACE total score (cognitive function) for all patients.The pink box highlights the crucial CG-SR discrepancy for AMI Total score as well as the three subscales. Values in the cells and background colour indicate Spearman’s ρ correlation coefficients. Correlations that did not survive correction for multiple comparisons are left blank. Bonferroni correction was applied for multiple comparison correction with α = 0.00045, computed based on 0.05 / total number of correlations computed.

AMI-CG scores also significantly correlated with self-reported anhedonia (measured by SHAPS total score using 4-point scoring system, ρ = 0.33, p < 0.001; **Figure 4**), which is considered a distinct construct. The total AMI-CG score in this study significantly correlated with self-reported depression (GDS; ρ = 0.25, *p* < 0.001). This remained significant even after removing the three apathy-related questions from the GDS (ρ = 0.24, *p* < 0.001).

These relationships between self-reported apathy, depression, and anhedonia were not unexpected, as these constructs often co-occur (**Figure 1**).^3^ Such relationships have been seen in large-scale studies of healthy people throughout lifespan and are consistent with the notion that apathy and anhedonia may share overlapping features.

Given that self-reported apathy scores were generally lower than caregiver-reported scores, we sought to determine whether this discrepancy reflects a systematic reporting bias (i.e., patients consistently rate their apathy lower while still being sensitive to variations in their own apathy levels) or whether patients and caregivers differ in their perception of apathy in a more complex way. Specifically, we tested whether self-reported apathy across different domains (Behavioural, Social, and Emotional) could still predict caregiver-reported total apathy scores. To quantify this potential agreement between self-reported and caregiver reported apathy, we conducted a multiple linear regression analysis. The self-reported scores on the three AMI subscales (Behavioural, Social, and Emotional) were entered as predictors of the caregiver-reported AMI total score. This model revealed that self-reported apathy, across all three domains, significantly predicted care-giver-reported total apathy (F(3, 331) = 12.97, *p* < 0.001, R2 = 0.141). Each of the three subscales contributed significantly to the model (t_behavioural_ = 2.75, p = 0.006; t_social_ = 2.93, p = 0.004; t_emotional_ = 2.90, p = 0.004). However, despite this “significant” prediction, the three self-report subscales accounted for only 14.1% of the variance in caregiver-reported total apathy scores.

This, combined with the moderate positive correlation found between AMI-SR and AMI-CG mentioned above, suggests that while there is partial agreement, there is also a clear divergence in how these two groups perceive and interpret patients’ apathetic behaviour. While a patient’s self-reported apathy provides some information about how their caregiver might perceive their apathy, it leaves most of the variance in caregiver report unexplained. This suggests that other factors, beyond the patient’s own perception of their apathy, influence how caregivers perceive and rate apathy.

Further investigation of the correlations within each diagnostic group revealed a striking pattern: self- and caregiver-reported AMI total scores were strongly and positively correlated in SVD (ρ = 0.63), PD (ρ = 0.57), SCD (ρ = 0.55), and AIE (ρ = 0.50) (all p ≤ 0.001). In AD, although self- and caregiver-reported scores showed a significant correlation, the relationship was moderate (ρ = 0.42, p = 0.002) (**Figure 5A** and **Figure 5B-D**). In contrast, no significant correlation was observed in bvFTD, SD, PDD, DLB, or MCI. Notably, bvFTD exhibited the most pronounced lack of agreement, with no correlation between self- and caregiver-reported scores (ρ = 0.12, p = 0.43). This lack of alignment in bvFTD was consistent across all three domains of apathy: Behavioral (ρ = 0.15, p = 0.34), Social (ρ = 0.25, p = 0.10), and Emotional (ρ = 0.07, p = 0.66).

**Figure 5:**
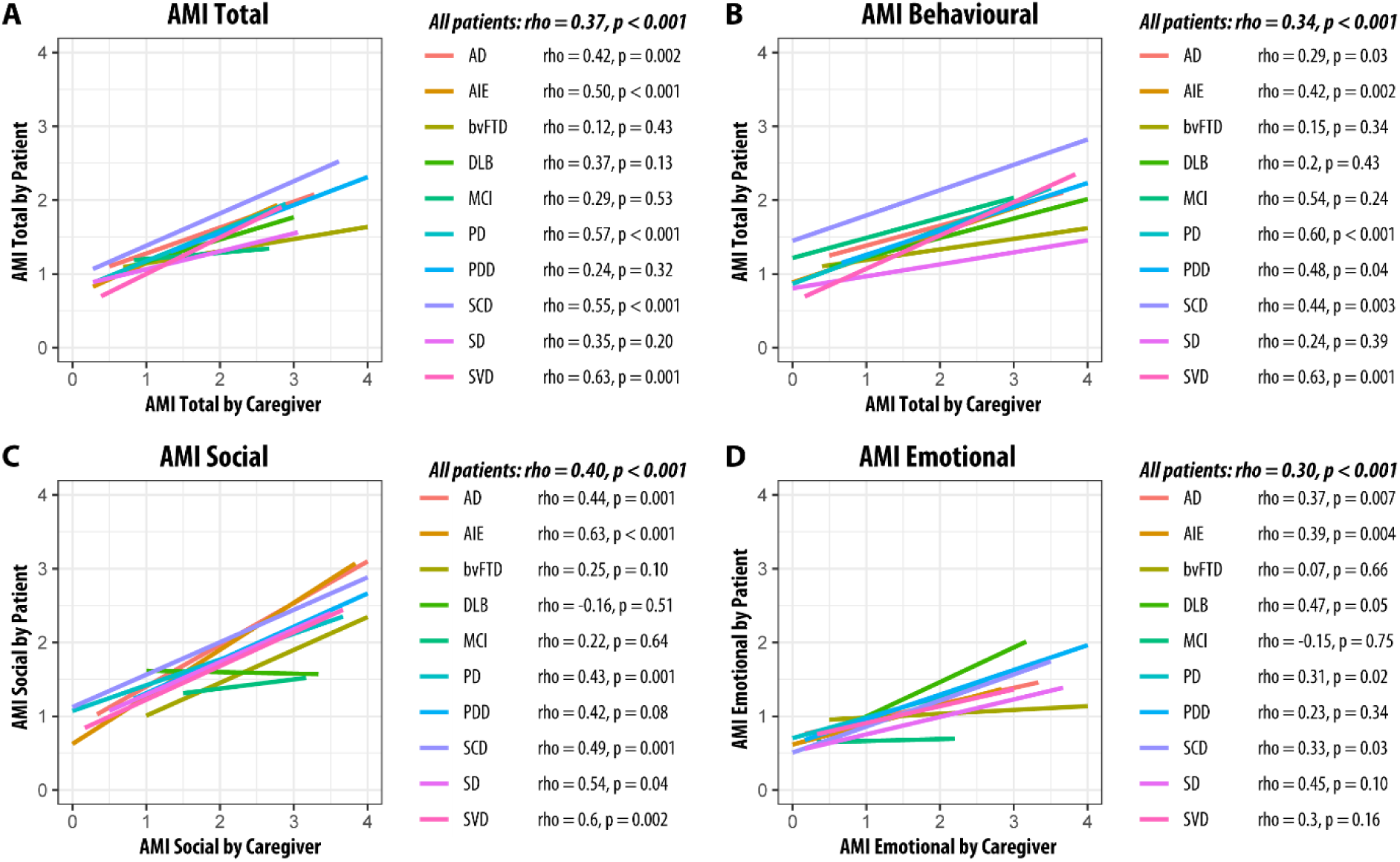
Correlation between self-reported and caregiver-reported apathy across patient groups. AMI-CG scores show moderate to strong correlations with their corresponding self-report scores in some, but not all, groups. (A) AMI Total score (B) AMI Behavioural subscale (C) AMI Social subscale (D) AMI Emotional subscale. Coloured lines indicate each cohort. Spearman’s ρ and p values are shown.

### Underestimation of apathy by patients with neurological conditions

To further understand this discrepancy, we examined the severity of apathy across different neurological conditions. Using established cut-offs on the AMI, we categorised apathy severity as none, moderate, or severe, based on both self-report and caregiver report (**Figure 6A** and **Figure 6B**; see detailed values in **Table 4**). (We define “moderate apathy” as apathy scores equal to or greater than one standard deviation above the mean AMI total score in a healthy population, i.e. AMI total score ≥ 1.91, and “severe apathy” as scores exceeding two standard deviations, i.e. AMI total score ≥ 2.38. This terminology was previously used in Ang et al.^41^ and we adopt the same classification here.) Of 335 patients, 14.0% self-reported moderate apathy, with a statistically similar prevalence based on caregiver report (17.9%, Chi^2^ = 1.6, p = 0.2). However, caregiver reports yielded a significantly higher prevalence of severe apathy across all conditions (Self-reported severe apathy 5.4% vs caregiver-reported severe apathy 23.9%, Chi^2^ = 44.5, p < 0.001). This underestimation by patients themselves was particularly pronounced in the bvFTD group, where self-reported severe apathy was only 9%, while caregiver reports indicated 58% with severe apathy (see more details in **Table 5**).

**Figure 6:**
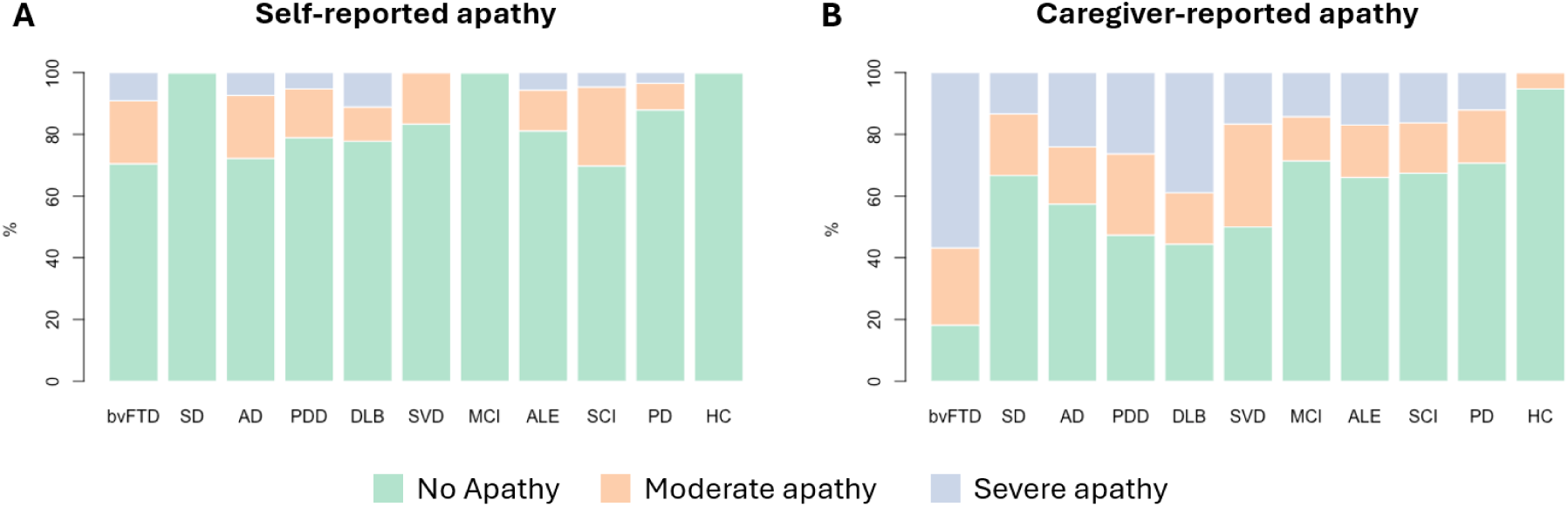
Prevalence of apathy based on patient and caregiver reports. The prevalence of apathy in different severity level is stacked as 100% for each group. The green portion of the bar represents no apathy (AMI Total < 1.91), orange indicates moderate apathy (AMI total score ≥ 1.91), and purple indicates severe apathy (AMI total score ≥ 2.38).

**Table 5:** Prevalence of different apathy severity based on patient and caregiver reports. Each cell shows the percentage values of No Apathy, Moderate Apathy and Severe Apathy. For example, based on self-report AMI total score, bvFTD’s data is 70/21/9, which means bvFTD had 70% no apathy, 21% moderate apathy and 9% severe apathy. SR = Self-report. CG = Caregiver-report.

| Group | Total |  | Behavioural Apathy |  | Social Apathy |  | Emotional Apathy |  |
| --- | --- | --- | --- | --- | --- | --- | --- | --- |
|  | SR | CG | SR | CG | SR | CG | SR | CF |
| bvFTD | 70/21/9 | 19/23/58 | 86/9/5 | 40/14/47 | 79/5/16 | 44/30/26 | 81/9/9 | 23/16/60 |
| SD | 100/0/0 | 62/25/12 | 94/6/0 | 88/6/6 | 75/25/0 | 75/12/12 | 100/0/0 | 75/6/19 |
| SVD | 83/17/0 | 50/33/17 | 88/8/4 | 67/25/8 | 83/17/0 | 71/25/4 | 96/0/4 | 58/25/17 |
| AIE | 81/13/6 | 66/17/17 | 87/8/6 | 70/21/9 | 74/21/6 | 72/19/9 | 96/2/2 | 77/15/8 |
| PD | 88/9/3 | 71/17/12 | 93/5/2 | 79/16/5 | 84/16/0 | 78/19/3 | 91/5/3 | 74/16/10 |
| PDD | 79/16/5 | 47/26/26 | 84/16/0 | 42/32/26 | 79/16/5 | 58/26/16 | 79/21/0 | 68/26/5 |
| DLB | 78/11/11 | 44/17/39 | 78/22/0 | 44/22/33 | 94/0/6 | 56/39/6 | 89/6/6 | 78/6/17 |
| AD | 72/20/7 | 57/19/24 | 80/20/0 | 67/17/17 | 69/22/9 | 69/24/7 | 87/9/4 | 70/7/22 |
| MCI | 100/0/0 | 71/14/14 | 86/14/0 | 57/43/0 | 100/0/0 | 71/29/0 | 100/0/0 | 71/29/0 |
| SCD | 70/26/5 | 67/16/16 | 74/21/5 | 86/9/5 | 65/33/2 | 65/26/9 | 91/7/2 | 79/14/7 |
| HC | 100/0/0 | 95/5/0 | 95/5/0 | 100/0/0 | 100/0/0 | 89/11/0 | 100/0/0 | 79/21/0 |

To explore the potential influence of cognitive impairment on the discrepancy, we ranked the patient groups by their average cognitive performance on the Addenbrooke’s Cognitive Examination III (ACE-III; **Figure 7A**). This analysis revealed a general trend for greater caregiver-self discrepancy in cohorts with poorer cognition (**Figure 7B**, see discrepancy in apathy subscales: **Figure 7C-E**) However, the discrepancy in bvFTD was significantly greater than in any other neurological condition, including AD, which had a similar level of cognitive impairment (no difference between their group ACE Total score: U=673.5, p = 0.46, ranksum r = 0.42). This finding suggests that factors beyond global cognitive impairment, such as lack of insight, may contribute to the underestimation of apathy in bvFTD.

**Figure 7:**
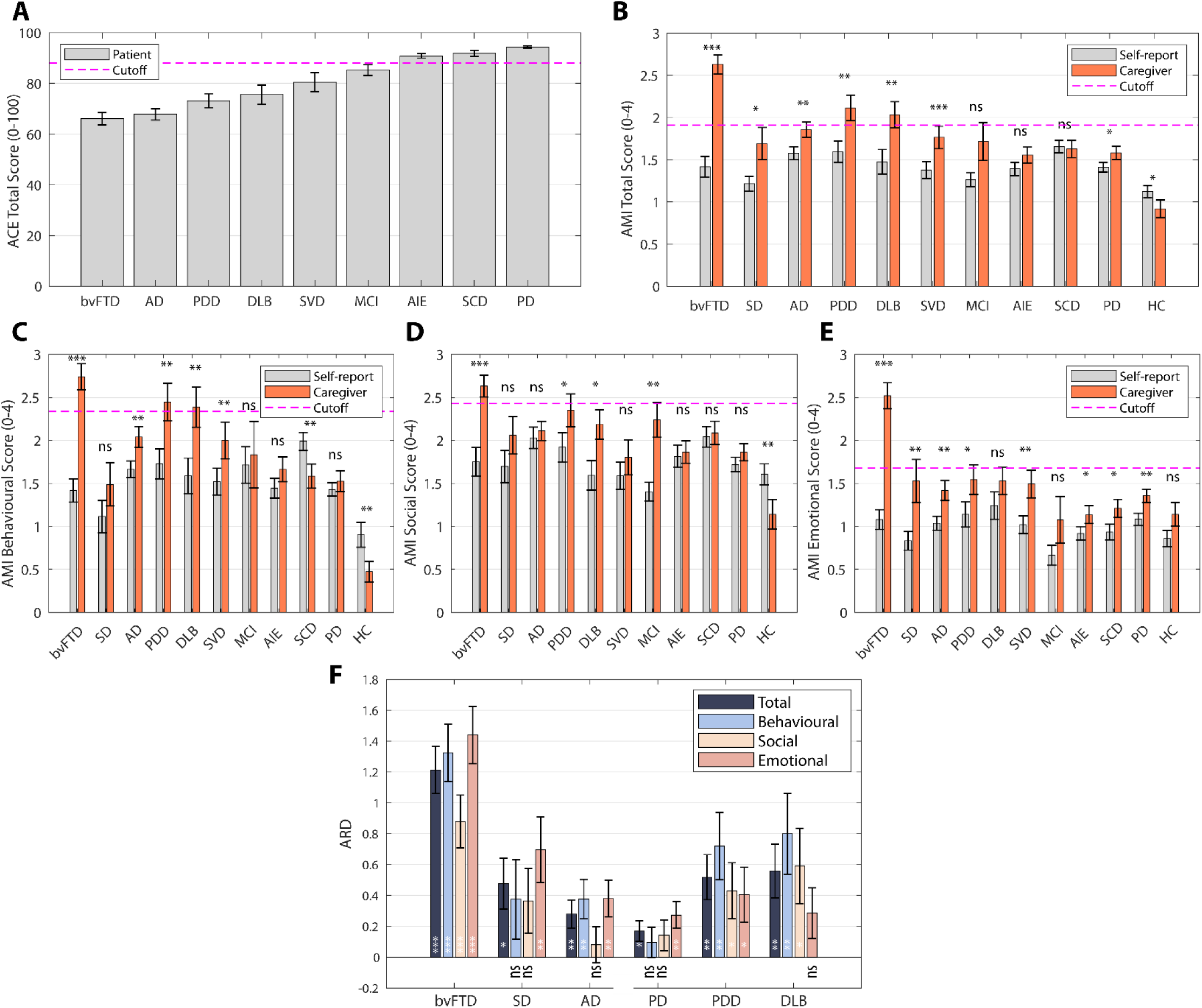
Caregiver-reported apathy and cognitive impairment. (A) Mean ACE total score for each cohort. The red horizontal dashed line indicates the cut-off for normal (88/100) ACE total score. Error bar indicates 1 standard error from mean (SEM). (B) Mean AMI total score for each cohort rated by patients themselves (grey bars) and their caregivers (orange bars). Results of paired t-tests comparing self- and caregiver-reported AMI total scores are shown above the paired bars for each cohort. (C-E) The same bar plots are shown for the subscales of AMI: Behavioural Apathy **(C)**, Social Apathy **(D)**, and Emotional Apathy **(E)**. **(F)** The apathy reporting discrepancy (ARD) for AMI Total, Behavioural, Social and Emotional subscore. In all plots, error bars indicate 1 SEM. Results of one-sample t-test against 0 is shown at the bottom of each bar. * p < 0.05, ** p < 0.01, *** p < 0.001. N.S. = not significant. p values were unadjusted.

Notably, the discrepancy between caregiver and self report was significant lager in PDD and DLB than PD (ARD in PDD vs PD: U=358.5, ranksum r = 0.35, p = 0.02; DLB vs PD: U=355, ranksum r = 0.32, p = 0.04; **Figure 7F**). In fact, the discrepancy in PD was mainly driven by a difference in emotional apathy, as there were no significant differences in apathy reporting in behavioural or social apathy (**Figure 7F**). Likewise, a significant discrepancy was observed in AD, but not in MCI or SCD (**Figure 7B**).

### Caregiver-Self Apathy Discrepancy and Cognition

To investigate the factors underlying the discrepancy between caregiver- and self-reported apathy, we calculated the Apathy Reporting Discrepancy (ARD) for each participant. ARD was defined as the difference between the AMI total score reported by the caregiver and the AMI total score reported by the patient (ARD = AMI_CG_ – AMI_SR_). Based on the subscale reported, we can also have ARD_Behavioural_ (i.e. discrepancy in AMI Behavioural subscale) and ARD_Social_, and ARD_Emotional_. Thus, ARD quantifies the extent to which caregivers perceive patients as more apathetic than the patients perceive themselves. **Figure 8A** presents a correlation matrix examining the relationships between ARD (total score and subscale scores) and various factors, including cognitive measures (ACE-III total and subdomain scores), depression (self-reported GDS depression-related items’ mean score, see Methods for rationale behind this choice), anhedonia (self-reported SHAPS total score), patient age, caregiver age, and education level.

**Figure 8:**
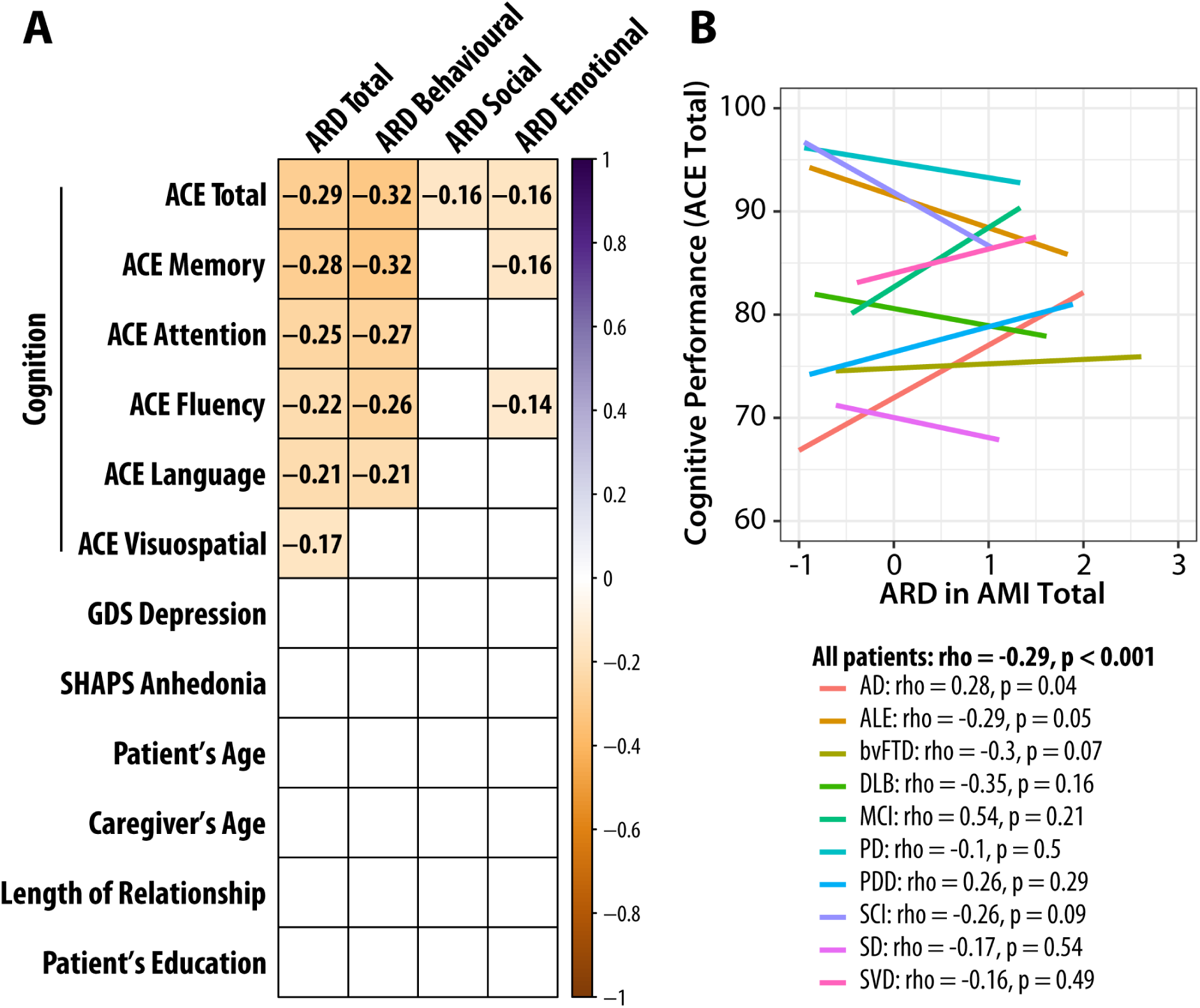
Correlation between Apathy Reporting Discrepancy (ARD) and cognitive measures. (A) Correlation matrix displaying the relationships between ARD, cognitive measures, depression, anhedonia, and demographic variables across all patients (N=314). Values in the cells and background colour indicate Spearman’s ρ correlation coefficients. Correlations that did not survive correction for multiple comparisons are left blank. Bonferroni correction was applied for multiple comparison correction with α = 0.001, computed based on 0.05 / total number of correlations computed. (B) Correlation between ARD Total and ACE Total (i.e., the top left corner in (A)) across patient groups. Coloured lines indicate each cohort. Spearman’s ρ and p values are shown.

Our analysis revealed that ARD could not be explained by patient depression (GDS Depression score, ρ = 0.007, p = 0.9, N = 295), anhedonia level (SHAPS Total score, ρ = -0.050, p = 0.4, N = 299), patient or caregiver age (ρ < 0.08, p > 0.2), length of the caregiver-patient relationship (ρ = 0.05, p = 0.4, N = 293), or patient education level (ρ = -0.12, p = 0.05, N = 259; this correlation did not survive correction for multiple comparisons).

Among the 314 patients with both ACE and AMI data^1^, ARD significantly negatively correlated with overall cognitive function (ACE total score; ρ = -0.29, p < 0.001) (**Figure 8B**). There are several possible explanations for this negative correlation between ARD and cognitive impairment: for example cognitively impaired patients may underestimate their own apathy, or fail to understand and respond to questionnaires meaningfully, or caregivers of patients with cognitive deficits endorse apathy items more commonly (possibly due to caregiver burden). The negative relationship between ARD and cognition was seen for all types of apathy, although with variation in the strength of correlation: Behavioural Apathy (ρ = -0.32, p < 0.001), with weak correlations with Social Apathy (ρ = -0.16, p = 0.005) and Emotional Apathy (ρ = -0.16, p = 0.004). ARD in Behavioural Apathy was associated with variation in all cognitive subdomains of the ACE: memory performance (ρ = -0.32, p < 0.001), and weakly associated with attention (ρ = -0.27, p < 0.001), verbal fluency (ρ = -0.26, p < 0.001), language (ρ = -0.21, p < 0.001), as well as visuospatial perception (ACE Visuospatial) (ρ = -0.16, p = 0.005). The correlation between cognitive function and ARD_Behavioural_ was strongest, significantly higher than both that with ARD_Social_ (z=-2.84, p=0.002) and ARD_Emotional_ (z=-2.44, p = 0.007).

To further examine the relationship between cognitive function and the discrepancy between caregiver and self-reported apathy (ARD), a stepwise multiple linear regression analysis was conducted using all five ACE subscores as predictors. Two models significantly predicted ARD across all patients: (1) Model 1 included only the memory subscore and was statistically significant, F(1,312) = 31.4, p < 0.001, explaining 9.2% of the variance in ARD (R² = 0.092). (2) Model 2 included both the memory and fluency subscores and was also statistically significant, F(2,311) = 18.9, p < 0.001, explaining 10.8% of the variance in ARD (R² = 0.108). These results suggest that memory function alone is a significant predictor of caregiver-self apathy discrepancy, but adding f luency as a predictor provides a modest increase in explained variance.

Furthermore, we re-ran the multiple linear regression on the subsample of 315 patients predicting total CG-apathy from AMI-SR subscales and cognition (ACE) data. We started with the model without cognition, which shows as previously that, the three self-reported AMI subscales explained 14.3% of the variance in caregiver-rated AMI total score (p < 0.001, F(3, 310) = 17.3, BIC = 687), with all three subscales contributing significantly (p ≤ 0.019, t ≥ 2.36).

To assess the contribution of specific cognitive domains, we compared this null model (three self-reported subscales) to a full model incorporating all ACE subscores (memory, attention, fluency, language, and visuospatial). Two models significantly improved upon the null model. The first model included, in addition to the three self-reported AMI subscales, the ACE fluency score. The model explained an additional 9.3% of the variance (total variance explained = 23.6%; F change (1,309) = 37.5, p < 0.001, BIC = 657.1). The substantial decrease in BIC (ΔBIC = 30.3) strongly suggests that the model incorporating the fluency performance provides a better fit to the data. The ACE fluency score contributed significantly (p<0.001, t = - 6.13), with a larger standardised coefficient value (-0.31) than all three self-reported AMI subscales (standardised coefficients ≤ 0.17). The second model included the ACE-III memory subscore in addition to fluency. Compared with the first model, the addition of ACE-III memory explained an additional 1.9% of the variance (F change (1, 308) = 7.95, p = 0.005, BIC = 654.9). Overall, these findings indicate that patients with greater cognitive impairment, particularly in verbal fluency and memory, exhibit a larger discrepancy between caregiver- and self-reported apathy.

## Discussion

This study revealed a significant discrepancy between self- and caregiver-reported apathy across multiple neurological disorders: bvFTD, SD, SVD, PD, PDD, DLB, AD, MCI, AIE and SCD (**Figures 5** & **7B**). This discrepancy was observed across all apathy subdomains—behavioral, social, and emotional (**Figure 7C-E**), and was more pronounced in conditions with greater cognitive impairment. For example, PDD and DLB demonstrated larger discrepancies than PD with normal cognitive performance. Similarly, the discrepancy was greater in bvFTD than AD, likely reflecting the more severe lack of insight associated with bvFTD despite similar scores on a global cognitive screening task (ACE). These findings underscore the critical importance of considering both patient and caregiver reports for accurately assessing apathy in neurological conditions.

An extensive literature has demonstrated that apathy is common in all types of dementia, with prevalence ranging from one thirds to almost 100%.^61^ For example, although apathy is commonly reported to be more severe in bvFTD compared to SD and AD,^15,62,63^ the reported prevalence in bvFTD still ranges widely from 50% to 90% across different studies.^15,43–46,62–69^ This variability may rise from the use of different screening tools, operational definitions of apathy, or thresholding approaches.^61^ Investigations using apathy-specfic scales like Starkstein’s Apathy Scale or Apathy Evaluation Scale (AES) generally report higher prevalence rates compared to those conducted with the Neuropsychiatric Inventory (NPI).^61,70^

In our study, self-reported data initially suggested a lower prevalence of apathy than expected. For instance, only 30% of bvFTD patients self-reported apathy on the AMI (**Table 3**), significantly below the 50–90% range reported in the literature using other scales.^61^ Further investigation revealed that this may be attributable to patient underreporting or the cut-off thresholds used. When caregiver-reported AMI scores were considered, 81% of bvFTD patients were apathetic, with 58% classified as severely apathetic (**Table 3**). Prevalence rates for other conditions in our study align closely with prior literature: 37% for SD, 50% for SVD^71^, 44% for AIE^72^ and 29% for PD^73^. As expected, apathy prevalence was higher in PDD (52%) and DLB (56%) than in PD.^74–76^ Consistent with prior findings, AD showed greater apathy prevalence (43%) compared to MCI (29%) and SCD (33%).^77,78^ In summary, these findings reveal the limitations of relying solely on self-report for diagnosing apathy, particularly in older adults and those with severe apathy.

Another important consideration is the use of cut-off thresholds in assessment tools for identifying apathy. Apathy exists along a continuous spectrum, varying in severity across one or more dimensions, rather than being a binary condition that is simply present or absent. However, it is common practice to define apathy based on a predetermined severity threshold. Thresholds derived from younger, healthy populations^41^, may not be suitable for older adults. Our unpublished AMI data from 1361 healthy adults over 50 years old suggest an adjusted cut-off (AMI Total ≥1.72). Using this adjustment, caregiver-reported prevalence increased to 91% in bvFTD and 79% in PDD. At the same time, such adjustment would not affect the apathy prevalence in the healthy control group. These findings underscore the need for standardisation in apathy assessment tools for more accurate and consistent evaluations in dementia research.

Previous reports have consistently shown that caregivers report higher levels of apathy in patients than patients report themselves.^15,21,24,27,30–32,42,79–81^ Without an objective measure of goal-directed behaviour, it remains unclear whether patients underreport their symptoms of caregivers overreport them. Individuals with apathy may perceive themselves as “content” and be less troubled by behavioural changes, whereas caregiver burden may heighten sensitivity to signs of apathy. Regardless of the underlying case, this discrepancy is striking—self-reported AMI scores for behavioural, social, and emotional domains together explained *only* 14.1% of the variance in caregiver-reported AMI scores. More importantly, we found that this discrepancy also varied significantly across disorders (**Figures 5** and **7**). In conditions such as SVD, PD, SCD, and AIE, self and caregiver ratings were strongly correlated, suggesting that the discrepancy reflected differences in severity rather than a fundamental disconnect. However, in conditions with greater cognitive impairment, such as PDD, DLB, AD and FTD, the correlation was weaker or even nonsignificant.

Here, patients’ cognitive impairment emerged as a key factor influencing the magnitude of the apathy reporting discrepancy. Specifically, across conditions greater cognitive impairment was associated with larger discrepancies. The positive relationship between apathy reporting discrepancy and patients’ cognitive impairment reported here cannot be solely attributed to a general caregiver reporting bias linked to increased caregiver burden. If caregiver burden were the primary driver, we would expect an overall inflation in caregiver-reported apathy scores equally across all apathy domains. However, as shown in **Figure 8A**, the relationship between ARD and cognition was specific to the behavioural AMI subscale, with the discrepancy being particularly pronounced in patients with greater impairments in memory, attention, and fluency. This suggests that cognitive deficits may selectively impact how patients perceive and report their own apathy, rather than caregivers simply overestimating apathy across the board.

Moreover, patients’ memory impairment and fluency deficits explained an additional 11.2% of the variance in caregiver-reported apathy. One possible explanation between this specific relation with fluency and memory deficits lies in the crucial role of executive function in goal-directed behaviour. In particular, cognitive fluency has been suggested to be essential for “option generation”—the ability to identify possible courses of action, which serves as a fundamental first step in motivated behaviour.^82^ Impaired fluency may restrict patients’ ability to spontaneously generate or consider different actions, resulting in inertia.^83^ Additionally, memory deficits could prevent patients from following through with plans or recalling past intentions, further reinforcing the caregiver’s perception of reduced motivation. Another possible explanation is that patients with greater memory impairment may simply forget their own levels of behavioral inactivity, leading them to underestimate their apathy. In contrast, caregivers may provide a more accurate account of apathetic episodes. Thus, rather than simply reflecting caregiver overestimation as proposed by previous studies^27,30,31,84^, the observed discrepancy between self- and caregiver-reported apathy may stem from distinct cognitive mechanisms, specifically deficits in memory and executive function, that shape how patients perceive and experience apathy across different neurological conditions.

Among conditions with similar cognitive screening total score, such as bvFTD and AD, the discrepancy was larger in bvFTD. (Note that the pattern of cognitive deficits over domains differ markedly in the ACE, only the total scores are comparable). This suggests the role of impaired insight in these patients on the apathy reporting discrepancy. In this group, the lack of agreement between self- and caregiver-reports was consistent across all three apathy domains, and the magnitude of the discrepancy was particularly striking. Cognitive impairment alone cannot fully explain this discrepancy, as AD patients, despite comparable cognitive deficits, demonstrated smaller discrepancies than bvFTD patients, consistent with previous report on bvFTD and AD.^21,62,85^ This pattern aligns with research on anosognosia in dementia, where discrepancies between self- and caregiver-reports of cognitive deficits have been linked to patients’ diminished self-awareness.^29,38,84,86^ In such cases, greater divergence between caregiver and patient ratings reflects more severe reductions in patients’ awareness.^84,87^

We also investigated whether patients’ depression or anhedonia could explain the apathy reporting discrepancy, because apathy, depression and anhedonia are very relevant in both healthy and patient population.^3^ Despite the frequent co-occurrence of apathy, depression, and anhedonia, our findings suggest that the underestimation of apathy by patients cannot simply be attributed to low mood or an inability to experience pleasure of patients themselves. This aligns with a previous study that found caregivers’ depression did not account for discrepancies in apathy reporting either.^30^ These findings reinforce that the discrepancy in apathy reporting is unlikely to result from negative emotional bias in patients or caregivers but rather reflects more complex factors, such as impaired patient awareness and cognitive impairment. In addition to these factors, caregiver burden could be another critical issue, though this was not investigated in the present study. Previous studies have shown that caregiver burden correlated with patients’ cognitive impairment and predicted the apathy reporting discrepancy.^21,30,31,47^ For instance, care-givers’ ratings of apathy increased over time in AD and MCI patients, even when patients’ self-reports remained stable.^31^

### Limitations

While our findings are robust, certain limitations should be considered. First, cohort sizes varied across conditions, potentially affecting statistical power. In particular, MCI group was small and the specific results for this group should be taken with caution. Second, we lacked an objective quantitative measure of apathy, or impaired insight, for all individuals. Third, the study did not directly measure caregivers’ depression or perceived burden, which could have influenced their ratings. Fourth, we used categorised (thresholded) classification of apathy based on supra/infra-threshold endorsements of apathy items, rather than continuous severity variables. This facilitates the comparison of groups, but we acknowledge that apathy within the “normal” range may still be relevant in the context of disease and its progression. Future research incorporating care-giver burden assessments might provide further understanding of contributions to apathy reporting discrepancies. Additionally, the proportion of caregivers in this study who were primary care-givers remains unclear.

## Conclusion

This study highlights the need to consider both patient and caregiver perspectives on apathy, especially for conditions with prominent cognitive impairment. To improve the recognition and management of apathy, and improve the understanding of the neurocognitive mechanisms of apathy, standardised assessment tools are required that are applicable to people with cognitive deficits. These will facilitate clinical interventions to benefit both patients and caregivers.

## Abbreviations

ARD: Apathy Reporting Discrepancy
AMI: Apathy-Motivation Index
AMI-CG: Apathy-Motivation Index Care-Giver Version
AMI-SR: Apathy-Motivation Index Self-Report Version
ACE-III: Addenbrooke’s Cognitive Examination-III
GDS: Geriatric Depression Scale Short Form
SHAPS: Snaith-Hamilton Pleasure Scale
bvFTD: Behavioural Variant Frontotemporal Dementia
SD: Semantic Dementia
SVD: Small Vessel Disease
AIE: Autoimmune Encepha-litis
PD: Parkinson’s Disease
PDD: Parkinson’s Disease Dementia
DLB: Dementia With Lewy Bodies
AD: Alzheimer’s Disease Dementia
MCI: Mild Cognitive Impairment
SCD: Subjective Cognitive Decline
HC: Healthy Controls.

## Footnotes

1 This includes ACE-R data from SD.

